# Pheno+: An Electronic Health Record-Integrated, Clinician-in-the-Loop Application for Efficient Phenotyping in Clinical Genomics

**DOI:** 10.64898/2026.11.02.26366992

**Authors:** Emerson L. Lebleu, Isabelle B. Cooperstein, Sabrina Malone Jenkins, Pinar Bayrak-Toydemir, Rong Mao, Alistair Ward, Tony Di Sera, Chelsea M. Solorzano, Tanner Ellsworth, Kelsey Simek, Peter Taber, Phillip Warner, Kensaku Kawamoto, Gabor T. Marth, Martin Tristani-Firouzi

**Author notes:** Corresponding author: Martin Tristani-Firouzi.

## Abstract

**Objective:** To evaluate the efficiency and diagnostic utility of Pheno+, an electronic health record (EHR)-integrated application that streamlines the generation of structured phenotypes for genomic testing and analysis.

**Materials and Methods:** We developed Pheno+ as a Substitutable Medical Applications and Reusable Technologies on Fast Healthcare Interoperability Resources (SMART on FHIR) application embedded within the University of Utah Health EHR. Pheno+ applies ClinPhen to clinician-selected clinical notes to identify candidate Human Phenotype Ontology (HPO) terms, which clinicians review and curate within the EHR. We conducted an eight-case crossover evaluation comparing workflow time between Pheno+ and a standard workflow that involved physician chart review and molecular geneticist-led HPO term selection. In a separate cohort of 17 patients with confirmed monogenic diagnoses, we assessed the diagnostic utility of Pheno+-derived HPO terms by evaluating Exomiser gene prioritization performance using these terms.

**Results:** Pheno+ reduced mean total workflow time by 66%, from 31.1 to 10.5 minutes (p < 0.001). Incorporating Pheno+-derived HPO terms improved Exomiser’s ability to rank the causative gene first, from 11 of 17 cases (64.7%) without phenotype input to 17 of 17 cases (100%) with phenotype input (p = 0.031).

**Discussion and Conclusion:** Pheno+ reduced the time and personnel required to generate clinician-curated HPO term lists while producing phenotypes that improved causative gene prioritization. These findings support EHR-integrated, clinician-in-the-loop phenotyping as an efficient approach to generating diagnostically informative phenotype data for clinical genomic workflows.

## INTRODUCTION

Genomic diagnosis of rare disease relies on integrating multiple sources of evidence to distinguish a small number of causative variants from the large background of genomic variation present in every individual. Variant prioritization tools, such as Exomiser [1], combine population allele frequencies [2], functional prediction scores [3–5], inheritance patterns, and patient phenotype data to rank candidate variants. Among these, patient phenotype data plays a central role by helping determine whether a candidate variant is consistent with the patient’s clinical presentation, rather than simply rare or predicted to be damaging [6]. To make this phenotypic information usable computationally, clinical phenotypes are typically encoded using the Human Phenotype Ontology (HPO) [7, 8], the standard vocabulary that supports phenotype-driven variant prioritization and diagnosis in rare diseases [9, 10].

Despite their diagnostic value, structured HPO annotations are rarely generated during routine clinical care. Instead, patient presentations are documented as free-text narratives distributed across numerous electronic health record (EHR) notes authored by multiple providers. Although these narratives contain rich phenotypic information, they cannot be readily used by computational diagnostic tools that require structured terms. Manually extracting and annotating phenotypic features from clinical notes is time-intensive and requires familiarity with HPO terminology that many providers lack, adding further burden to already demanding clinical workflows. Fully automated extraction tools can reduce this burden, yet natural language processing (NLP) methods remain imperfect and can introduce errors that affect diagnostic accuracy [11, 12]. Therefore, clinician review remains important for identifying and correcting errors in automatically extracted terms. Approaches that preserve this clinician oversight may help translate narrative clinical documentation into computable phenotype representations without sacrificing accuracy.

As genomic sequencing is increasingly integrated into clinical care, generating structured phenotype data at scale represents an important workflow challenge. Automated extraction of phenotypic information from clinical documentation has therefore been explored as a strategy to reduce the manual effort required for genomic analysis and to enable structured phenotyping across larger patient populations [13, 14]. Our experience implementing genomic testing in the neonatal intensive care unit (NICU) through the Utah NeoSeq program highlighted this challenge in a clinical setting. NeoSeq provides rapid genomic evaluation for critically ill neonates with suspected genetic disorders within the University of Utah healthcare system [15]. Generating a comprehensive HPO term list is a required step in this evaluation, supporting downstream variant prioritization and diagnostic interpretation. The standard workflow for producing this list is a two-step manual process: a physician reviews clinical documentation and writes a narrative summary of the patient’s clinical phenotypes, which a laboratory molecular geneticist then reviews to generate a list of HPO terms. This sequential process requires the involvement of both the physician and the molecular geneticist and introduces an additional step between clinical assessment and structured phenotype generation.

Widespread adoption of the Substitutable Medical Applications and Reusable Technologies (SMART) on Fast Healthcare Interoperability Resources (FHIR) framework [16] has enabled secure integration of third-party applications directly within EHR systems via standardized FHIR interfaces. By enabling applications to access clinical data through standardized interfaces, SMART on FHIR provides an opportunity to integrate informatics tools into existing clinical workflows without requiring clinicians to leave the EHR.

To address the challenges of adopting structured phenotyping within routine patient care, we developed Pheno+, a SMART on FHIR application that retrieves relevant clinical notes, translates narrative findings into candidate HPO terms, and enables clinicians to review and refine phenotype annotations directly within the EHR. Here, we describe the implementation of Pheno+ and evaluate its impact on the efficiency of clinical phenotyping workflows and the utility of the resulting HPO terms for downstream genomic analyses.

## METHODS

### System architecture and implementation

Pheno+ was developed as a web-based application using the Vue.js front-end framework and the Vite development and build toolkit. The application interfaces with electronic health record (EHR) data through standardized SMART on FHIR protocols [16]. Pheno+ is hosted entirely within the University of Utah’s protected server infrastructure in compliance with institutional policies governing protected health information (PHI).

### EHR integration and SMART on FHIR configuration

Pheno+ was implemented as a SMART on FHIR application integrated into the Epic EHR environment. Authentication and authorization are managed using the SMART on FHIR JavaScript client library, fhirclient.js [17], which supports OAuth 2.0-based access to EHR resources. Following authorization, the application receives a FHIR client instance with permissions defined by the institutional EHR configuration. Pheno+ uses this client to retrieve patient clinical documentation and associated metadata through available FHIR endpoints. This integration allows clinicians to launch Pheno+ directly from the patient chart without leaving the EHR, supporting its use within existing clinical workflows.

### Phenotype extraction pipeline

To generate candidate HPO terms from clinical notes, we integrated ClinPhen [13], an NLP tool for extracting phenotype terms from unstructured clinical text. ClinPhen was deployed as a web-accessible service that interfaces with the Pheno+ backend. Clinical notes selected by the clinician are processed by ClinPhen, which returns candidate HPO terms for review (**Fig. 1**). For each candidate term, Pheno+ displays the corresponding clinical context and frequency of occurrence within the selected notes. Clinicians can remove extracted terms that do not accurately represent the patient’s phenotype and manually add relevant terms that ClinPhen did not identify. The final clinician-curated HPO term list can then be copied for downstream use.

**Figure 1:**
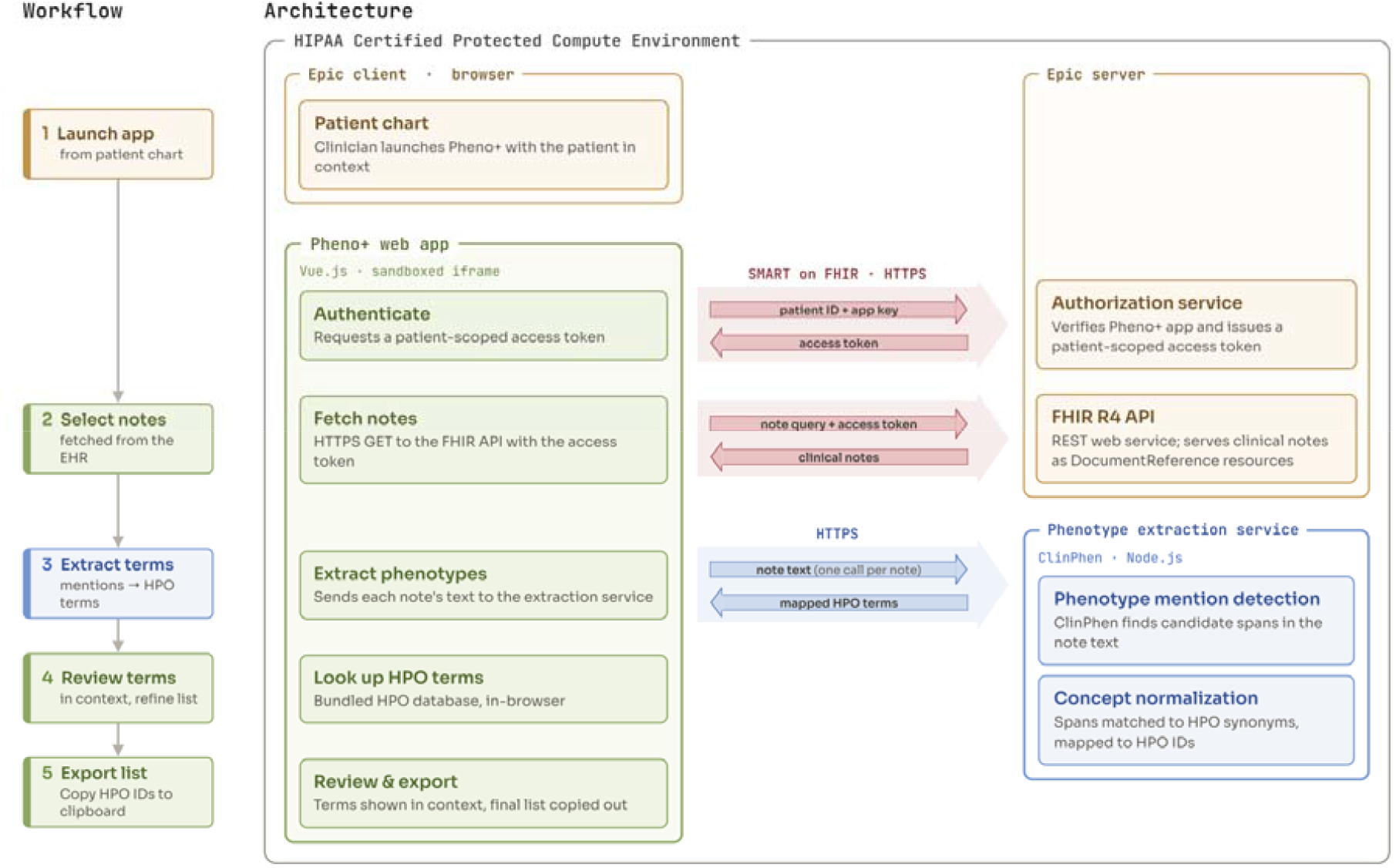
Pheno+ system architecture and data flow. The clinician launches Pheno+ from the electronic health record (EHR) using SMART on FHIR, which authenticates and authorizes the application via the FHIR server. Once authorized, Pheno+ retrieves clinician-selected clinical notes and submits their content to the ClinPhen server-side endpoint for phenotype extraction. ClinPhen returns candidate Human Phenotype Ontology (HPO) terms, which are displayed in the Pheno+ interface for clinician review and curation.

### Clinical workflow comparison and evaluation design

Two workflows for generating HPO term lists were compared: the existing NeoSeq workflow (“standard”) and the Pheno+ workflow. In the standard workflow, an attending neonatologist reviewed the patient’s clinical documentation and produced a brief narrative summary listing the key phenotypic features. A molecular geneticist then reviewed this summary and translated the clinical findings into a list of HPO terms to accompany genetic testing. This workflow required two professionals working in sequence.

In the Pheno+ workflow, the attending neonatologist independently generated the HPO term list without molecular geneticist involvement. The physician selected relevant clinical notes within the EHR for processing by Pheno+, which extracted candidate HPO terms from the selected documentation. The physician then reviewed and curated this list by examining the supporting clinical context, removing inaccurate terms, and adding relevant terms as needed to accurately represent the patient’s presentation.

To compare these workflows, eight cases from the NeoSeq program [15] were randomly assigned to one of two workflow sequences (n=4 per sequence) in a crossover design. In the standard-first sequence, HPO terms were generated using the standard workflow, followed by the Pheno+ workflow two weeks later. In the Pheno+-first sequence, the order was reversed. The two-week interval was intended to reduce potential recall bias, as the same physician performed the clinical review steps in both workflows.

For each case, we recorded the time required to complete each workflow step and the total number of HPO terms generated. Total workflow time, physician time, and HPO term counts were compared between workflows using paired t-tests. Potential order effects were assessed by comparing outcomes between the standard-first and Pheno+-first workflow sequences.

All physician review steps were performed by a single attending neonatologist. The second stage of the standard workflow was performed by two molecular geneticists, with cases evenly distributed between them.

### Diagnostic utility of Pheno+-derived phenotype terms

To evaluate the utility of Pheno+-derived HPO terms for downstream analysis, we identified a separate cohort of 17 NeoSeq patients with confirmed monogenic diagnoses. For each patient, the same attending neonatologist who participated in the workflow evaluation used Pheno+ to generate an HPO term list from clinical notes documented prior to the patient’s genetic diagnosis, thereby minimizing the risk that term selection was influenced by knowledge of the diagnostic result. These HPO term lists, together with each patient’s family-level variant call data, were provided to Exomiser (v.14.0.0) [1] to prioritize candidate diagnostic genes. To assess the contribution of Pheno+-derived phenotype information, Exomiser was also run using the same variant data without phenotype input. For each patient and condition, we recorded the rank of the known causative gene among the candidates returned by Exomiser. The proportion of cases in which the causative gene ranked first was compared between conditions using McNemar’s exact test.

## RESULTS

### System implementation and workflow

Pheno+ was developed and deployed over a five-month period, encompassing EHR integration, user interface development, and workflow testing, demonstrating that a clinically integrated phenotyping tool can be built using existing technologies without requiring extensive development resources. Pheno+ enables semi-automated extraction and clinician-guided curation of HPO terms from clinical documentation through an interface accessible directly from the EHR.

Pheno+ launches within the context of a patient chart (**Fig. 2**). Upon launch, the application retrieves available clinical notes through FHIR endpoints and presents them in a selectable list. Patient charts may contain multiple types of documentation, including admission notes, progress notes, subspecialty consultation reports, and discharge summaries, not all of which are relevant to the suspected genetic condition. The Pheno+ workflow proceeds through the following steps:

1. The clinician browses and selects clinical notes that are likely to contain phenotypic details relevant to the patient’s suspected genetic condition.
2. Selected notes are processed by ClinPhen to extract candidate HPO terms.
3. Extracted HPO terms are displayed in a table alongside contextual excerpts from the source note and their frequency of occurrence.
4. The clinician reviews and curates the candidate terms, removing those that do not accurately represent the patient’s phenotype and manually adding relevant terms not identified by ClinPhen.
5. The final clinician-curated list of HPO terms is copied for downstream genomic analysis.

**Figure 2:**
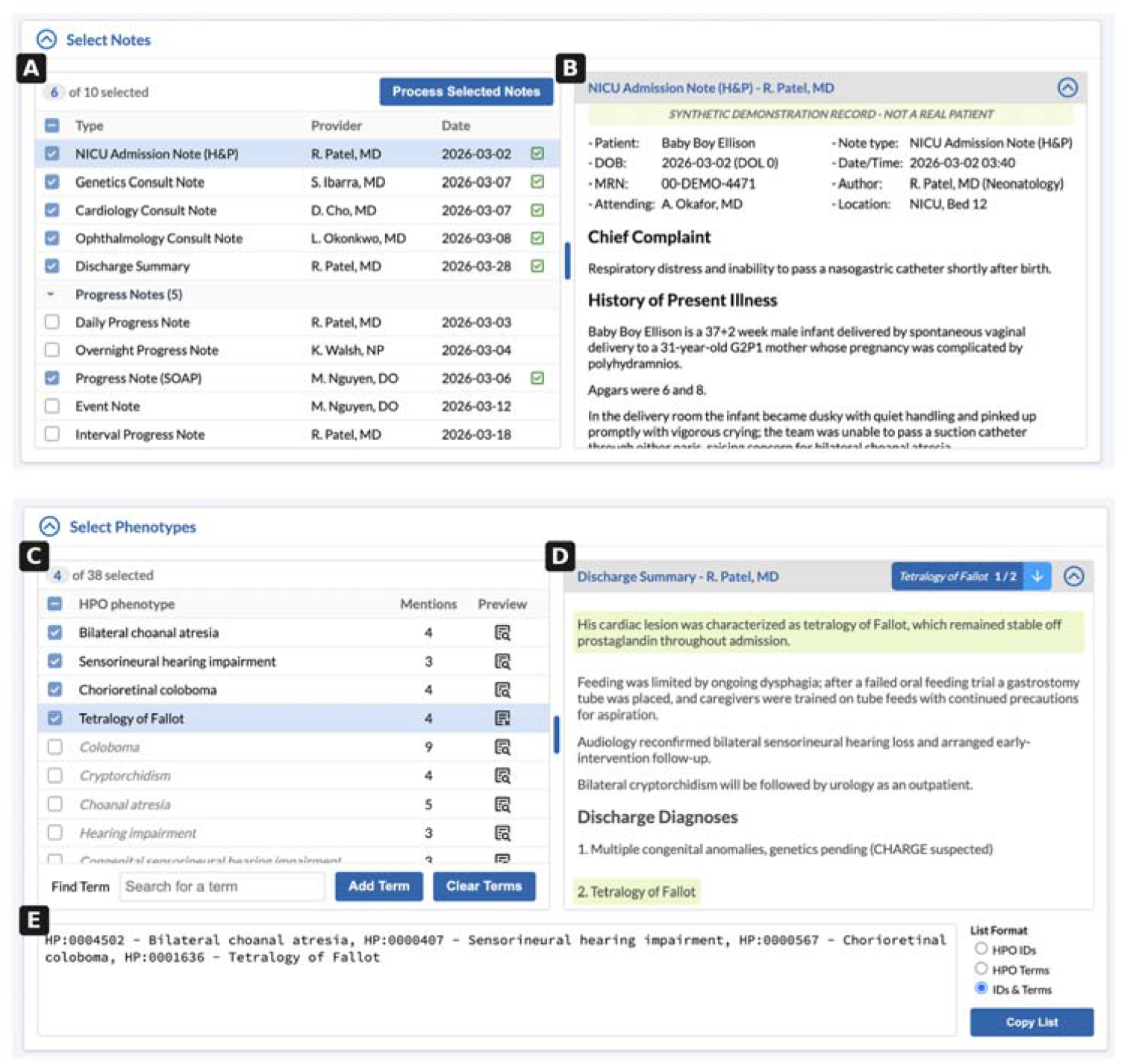
Pheno+ user interface. The interface is shown using a synthetic patient record. Clinicians select relevant clinical notes from the patient’s chart (A) and view note content directly within the application (B). Selected notes are processed by ClinPhen to generate a table of candidate HPO terms (C), with contextual excerpts from the source notes available for each term to support clinician review (D). Clinicians review and curate the extracted terms, removing those that do not accurately represent the patient’s phenotype and retaining relevant terms. The final clinician-curated HPO term list is displayed in a format that can be copied for downstream genomic analysis (E).

Pheno+ does not currently persist session data between launches. Consequently, note selections and curated HPO terms must be regenerated if the application is closed before the final list is copied.

### Workflow efficiency and phenotype yield in the eight-case crossover evaluation

Pheno+ was designed to enable HPO terms to be generated by the treating physician, who has firsthand knowledge of the patient, while minimizing the time required. We evaluated the efficiency of Pheno+ relative to the standard workflow across eight NeoSeq cases using a crossover design (Methods). The Pheno+ workflow required an average of 10.5 minutes of staff time per case, compared with 31.1 minutes for the standard workflow, representing a 66% reduction in workflow time (paired t-test, p = 7×10^−5^; **Table 1**). Notably, the physician review and phenotype summarization step of the standard workflow alone required an average of 19.8 minutes, exceeding the average total time required to complete the entire Pheno+ workflow (10.5 minutes; paired t-test, p = 0.00075). Whereas the standard workflow required the sequential involvement of a physician and a molecular geneticist, Pheno+ enabled the physician to independently generate the HPO term list.

**Table 1.**
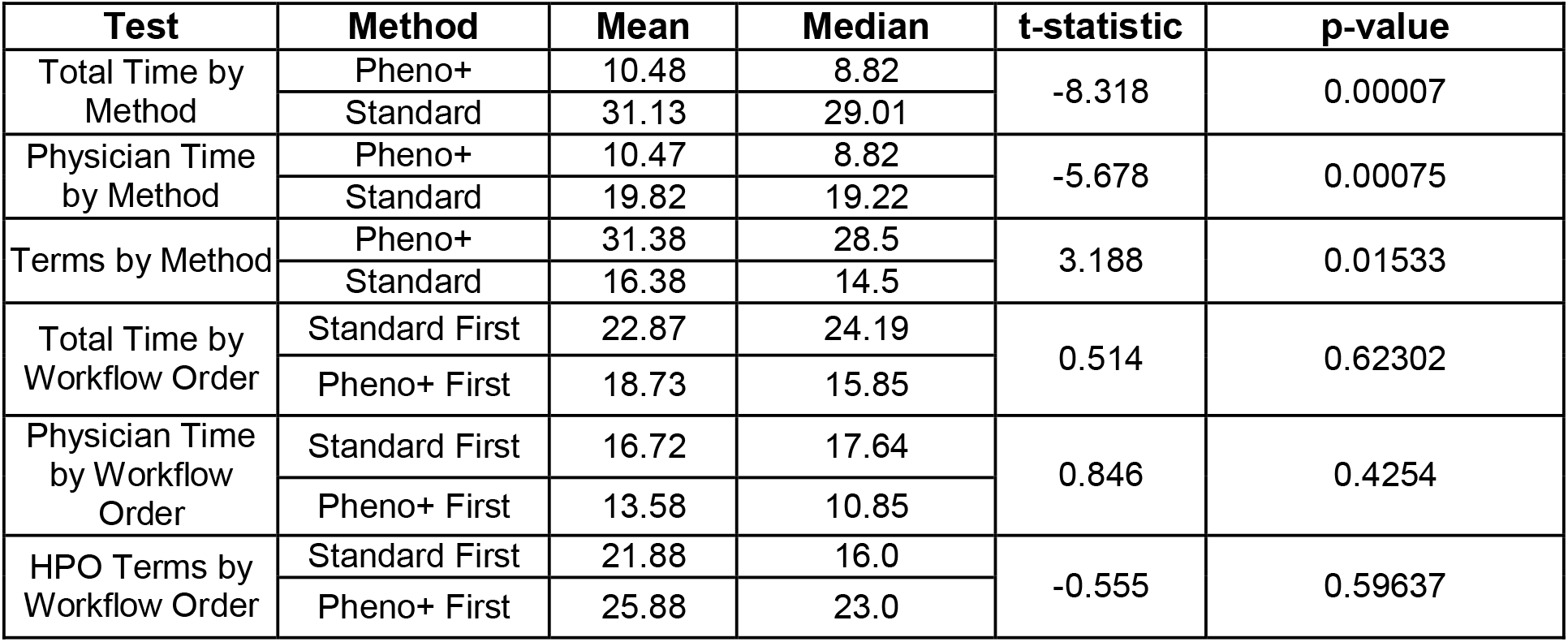
Summary of workflow time and phenotype term yield across the eight-case crossover evaluation.

Pheno+ also produced significantly more HPO terms than the standard workflow, with an average of 31.4 terms per case compared with 16.4 terms (paired t-test; p = 0.0153; **Table 1**). Before clinician curation, ClinPhen generated larger candidate lists, from which clinicians removed an average of 21.9 terms during review.

Comparisons between the standard-first and Pheno+-first sequences found no statistically significant differences in total workflow time (p = 0.623), physician time (p = 0.425), or HPO term count (p = 0.596), providing no evidence of the crossover design introducing a systematic bias.

Total time includes both physician and molecular geneticist time for the standard workflow and physician time alone for the Pheno+ workflow. Total time, physician-only time, and number of HPO terms generated are compared between the Pheno+ and standard workflows (top three rows) and by workflow order to assess potential order effects in the crossover design (bottom three rows). For workflow order comparisons, cases completed either the standard workflow followed by Pheno+ or Pheno+ followed by the standard workflow, with a two-week interval between methods. Values reflect mean and median per case; comparisons were performed using paired t-tests.

### Diagnostic utility of Pheno+-derived phenotype terms

To evaluate the utility of Pheno+-derived HPO terms for diagnostic gene prioritization, we analyzed a separate cohort of 17 NeoSeq cases with confirmed monogenic diagnoses (Methods). For each case, Exomiser was run using family-level variant data with and without Pheno+-derived HPO terms. With Pheno+-derived HPO terms, Exomiser ranked the known causative gene as the top candidate in all 17 cases (100%; **Table 2**), compared with only 11 of 17 cases (64.7%) when Exomiser was run without phenotype input. In the remaining six cases, the causative gene ranked between 8th and 19th when phenotype information was omitted. This difference in gene ranking performance between the two conditions was statistically significant (McNemar’s exact test, p = 0.031).

**Table 2.** Exomiser gene ranking results with and without Pheno+-derived phenotype terms.

| **Case** | **Rank<br>(Pheno+ terms)** | **Rank<br>(No phenotype input)** |
|---|---|---|
| **1** | 1 | 10 |
| **2** | 1 | 10 |
| **3** | 1 | 1 |
| **4** | 1 | 1 |
| **5** | 1 | 1 |
| **6** | 1 | 1 |
| **7** | 1 | 1 |
| **8** | 1 | 1 |
| **9** | 1 | 19 |
| **10** | 1 | 1 |
| **11** | 1 | 1 |
| **12** | 1 | 1 |
| **13** | 1 | 15 |
| **14** | 1 | 10 |
| **15** | 1 | 8 |
| **16** | 1 | 1 |
| **17** | 1 | 1 |

For each of 17 patients with a confirmed monogenic diagnosis, Exomiser was run on family-level variant data together with either Pheno+-derived HPO terms or no phenotype input. The table shows the rank of the known causative gene among all candidates returned by Exomiser under each condition.

## DISCUSSION

Pheno+ significantly reduced the time and personnel required to generate HPO term lists in our NICU workflow by enabling the treating physician to complete phenotype extraction and curation without requiring subsequent review by a molecular geneticist. Because phenotype curation is performed by the treating clinician with firsthand knowledge of the patient, Pheno+ enables direct application of clinical judgment rather than relying on a secondhand interpretation of a written summary.

Beyond improving workflow efficiency, it is essential that Pheno+ generate phenotype terms that are informative for downstream genomic analyses. Exomiser ranked the causative gene first in every case evaluated using Pheno+-derived HPO terms, demonstrating that Pheno+ can efficiently generate clinician-curated phenotype terms that provide meaningful information for downstream analyses. However, automated phenotype extraction does not eliminate the need for clinician oversight, as candidate terms must still be reviewed for accuracy and clinical relevance. Integrating this review process into routine clinical practice may require adjustments to existing workflows to accommodate clinician curation of automatically generated phenotypes.

Although ClinPhen was selected for this implementation, the Pheno+ system architecture is designed to accommodate alternative phenotype extraction tools with minimal modification, allowing the application to adapt as NLP and large language model approaches evolve. This flexibility is particularly valuable given the well-recognized challenges of extracting phenotypes from unstructured clinical documentation. NLP systems must accurately interpret clinical context, including distinguishing patient phenotypes from family history (e.g., maternal disease history), handling negation and uncertainty, and avoiding extraction of incidental or non-patient findings as true clinical phenotypes [11, 12]. The clinician review step in Pheno+ is designed to identify and correct such errors, but a more accurate extraction backend would further reduce this curation burden.

## CONCLUSIONS

Pheno+ addresses a key bottleneck in clinical genomic testing by enabling clinicians to generate curated HPO term lists directly within the EHR. Pheno+ reduced total workflow time, eliminated the need for a separate molecular geneticist review step, and generated phenotype terms that provided meaningful information for downstream diagnostic gene prioritization. These findings support clinician-in-the-loop structured phenotyping as a practical strategy for improving the efficiency and scalability of clinical genomic workflows. Larger, multisite studies are needed to evaluate generalizability across clinical settings, users, and patient populations.

## Code Availability

Pheno+ is freely available with source code and installation instructions hosted at https://github.com/iobio/pheno-plus.

## Data Availability

The datasets used and/or analyzed during the current study are available from the corresponding author on reasonable request.

## Ethics approval

The research was reviewed by the University of Utah’s Institutional Review Board and received an exemption determination in accordance with category 2 (IRB #00165986).

## Author Contributions

E.L.L., S.M.J., G.T.M., A.W., K.K., and M.T.F. contributed to the conception and design of the study. E.L.L. developed the Pheno+ application, performed the workflow efficiency analyses, and drafted the original manuscript. S.M.J., C.M.S., T.E., and K.S. contributed to clinical study coordination, patient data collection, and workflow evaluation. P.B.T. and R.M. performed molecular geneticist review and HPO term generation for the standard clinical workflow. I.B.C. performed the Exomiser analyses and contributed to manuscript revision. A.W., P.T., P.W., and K.K. contributed to software development and EHR integration. T.D.S. generated Figures 1 and 2. G.T.M. and M.T.F. supervised the study and contributed to the interpretation of the results. All authors reviewed the final manuscript.

